## Supplemental tables, figures, and analyses for "Evaluation of FluSight influenza forecasting in the 2021-22 and 2022-23 seasons with a new target laboratory-confirmed influenza hospitalizations"

Mathis et al.

**Supplemental Materials**

Table S1: Model information

Figure S1: Distribution of relative WIS across subnational jurisdictions

Supplemental Analysis 1. Backfill/Revision of hospital admission data

Supplemental Analysis 2: Performance of log-transformed forecasts

Supplemental Analysis 3. Performance of national forecasts

Table S1: Types of models included in analysis

| Model Name | Type | Methods | Additional data used* | Repo_url |
|---|---|---|---|---|
| **CEID-Walk** (2021-22) | Statistical | Random walk model without drift. The last few observations of a target time series are used to estimate the variance in step size of the random walk. | | |
| **CEPH-Rtrend_fluH** (2022-23) | Statistical | Renewal equation based on a Bayesian estimation of R(t) from hospitalization data. | | https://publichealth.indiana.edu/research/faculty-directory/profile.html?user=majelli https://github.com/paulocv/Rtrend-fluH |
| **CMU-TimeSeries** (2021-22) | TS regression Statistical | A quantile autoregression fit using influenza-related hospitalizations and doctors visits, jointly trained across locations using the most recently available 21 days of data. | HHS field estimated percentage of outpatient doctors visits with confirmed influenza | https://github.com/cmu-delphi/flu-hosp-forecast/ |
| **CMU-TimeSeries** (2022-23) | TS regression Statistical | A quantile autoregression fit using influenza-related hospitalizations and doctors visits, jointly trained across locations using the most recently available 21 days of data.ensembled with another quantile autoregression model with a longer time window and adjustments for data latency. | HHS field estimated percentage of outpatient doctors visits with confirmed influenza | https://github.com/cmu-delphi/flu-hosp-forecast/ |
| **CU-ensemble** (2021-22,2022-23) | Ensemble | An inverse-WIS weighted ensemble of three models – an SEIRS compartmental model with EAKF, an ARIMA model, and a baseline model of random walk with drift. | "Daily climatology of specific humidity for each state compiled from the National Land Data Assimilation System | |

| | | | | |
|---|---|---|---|---|
| | | | (NLDAS) project-2 data set" | |
| **GH-Flusight** (2021-22) | Ensemble | National and State-Level model, providing daily forecast predictions of flu patients admitted to the hospital, providing forecasts for next four weeks. Model was an ensemble development | | https://github.com/gh-ai-solu/flusight-21-22 |
| **GT-FluFNP** (2021-22,2022-23) | Statistical | Data-driven approach based on probabilistic non-parametric neural Gaussian processes using syndromic, clinical, demographic, and mobility data. | Demographic, Google mobility, Facebook symptomatic data, JHU-CSSE, COVID-Net | https://github.com/AdityaLab/EpiFNP |
| **IEM_Health-FluProject** (2021-22) | Statistical | State- and national-level SEIR model projections for weekly incident confirmed flu hospitalizations by using MCMC to fit actual hospitalizations observed. | | |
| **JHU_IDD-CovidSP** (2022-23) | Mechanistic | State-level metapopulation SEIR model with inter-state human mobility, explicit modeling of age-structure, vaccination, immunity waning, and Flu types A and B. The disease transmission is changed over time by non-pharmaceutical/behavioral interventions and humidity-driven seasonality. | | https://www.flepimop.org/ |
| **LosAlamos_NAU-Cmodel_Flu** (2021-22) | Mechanistic Statistical | Seasonal compartmental model to perform Bayesian analysis on hospitalization data available for each state. | | https://github.com/computationalUncertaintyLab/2022Flu_LucompUncertLab |
| **LucompUncertLab-TEVA** (2021-22) | Statistical Ensemble | The four VAR models submitted by LU are combined by Thompson sampling with weights assigned at the location level and proportional to the number of times a model has the lowest WIS in that location. | Four component models submitted by Lehigh University | https://github.com/computationalUncertaintyLab/2022Flu_LucompUncertLab |
| **LucompUncertLab-VAR2** (2021-22) | Statistical | VAR(2) model that groups state-level hospitalization trajectories by HHS region and is fit with stan. | | https://github.com/computationalUncertaintyLab/2022Flu_LucompUncertLab |
| **LucompUncertLab-VAR2_plusCOVID** (2021-22) | Statistical | VAR(2) model that groups state-level hospitalization trajectories due to influenza and COVID-19 and is fit with stan. | HHS state-level confirmed COVID-19 hospitalizations aggregated to weeks. | https://github.com/computationalUncertaintyLab/2022Flu_LucompUncertLab |

| | | | | |
|---|---|---|---|---|
| **LucompUncertLab-VAR2K** (2021-22) | Statistical | VAR model that groups state-level hospitalization trajectories due to influenza and COVID by HHS region, selects the number of lagged weeks between 1 to 5 weeks behind, and is fit with stan. | HHS state-level confirmed COVID-19 hospitalizations aggregated to weeks. | https://github.com/computationalUncertaintyLab/2022Flu_LucompUncertLab |
| **LucompUncertLab-VAR2K_plusCOVID** (2021-22) | Statistical | VAR model that groups state-level hospitalization trajectories due to influenza and COVID-19, probabilistically selects the number of lags from 1 to 5 weeks, and is fit with stan. | HHS state-level confirmed COVID-19 hospitalizations aggregated to weeks. | https://github.com/computationalUncertaintyLab/2022Flu_LucompUncertLab-VAR2 |
| **MIGHTE-Nsemble** (2022-23) | Machine-Learning Ensemble | Statistical Ensemble encompassing a set of network-based models, including Vector Autoregression and Lasso, as well as Exogenous Data models such as ARGO. | Influenza-related Google Searches retrieved from Google Trends. | |
| **MOBS-GLEAM_FLUH** (2021-22,2022-23) | Mechanistic Network | Metapopulation, age structured SLIR model, including local mobility and domestic airline traffic and school calendar. | HHS state-level confirmed COVID-19 hospitalizations aggregated to weeks. | https://www.gleamproject.org/flu-forecasting |
| **PSI-DICE** (2021-22,2022-23) | Mechanistic | A stochastic/deterministic, SIRH model that stratifies by disease severity and includes compartments for hospitalizations. Parameter posteriors are inferred using an MCMC procedure. | | |
| **Sgroup-RandomForest** (2021-22,2022-23) | Ensemble | Random Forest ensemble of the predictors generated from the Sgroup-SikJalpha submission, HHS data, and FluSurv-NET data. | FluSurv-NET | https://github.com/maa989/RandomForest-SikJalpha |
| **Sgroup-SlkJalpha** (2021-22) | Statistical | Discrete heterogeneous rate model where rates are learned using regression with a forgetting factor that weighs recently seen data higher. | | https://github.com/scc-usc/ReCOVER-COVID-19 |
| **SigSci-CREG** (2021-22,2022-23) | Statistical | Count regression model. | | https://github.com/signaturescience/fiphde |
| **SigSci-TSENS** (2021-22,2022-23) | TS ensemble | Ensemble of time series models. | | https://github.com/signaturescience/fiphde |
| **UGA_flucast-OKeeffe** (2022-23) | | | | https://thefoxlab.wordpress.com/ |
| **Umass-trends_ensemble** (2021-22,2022-23) | TS ensemble | Equally weighted ensemble of simple time-series baseline models. | | https://github.com/reichlab/flu-hosp-models-2021-2022 |
| **UT_FluCast-Voltaire** (2021-22) | Statistical | Rolling Bayesian taylor approximation on logarithm of cases implemented in stan. | HHS previous day admission influenza confirmed coverage | |

| | | | |
|---|---|---|---|
| **UVAFluX-Ensemble** (2021-22) | TS ensemble | Ensemble of multiple methods such as auto-regressive (AR) models with exogenous variables, ARIMA, Kalman filter and Long short-term memory (LSTM) models | |
| **UVAFluX-Ensemble** (2022-23) | TS ensemble | Ensemble of multiple methods such as auto-regressive (AR) models with exogenous variables, ARIMA, Kalman filter, Long short-term memory (LSTM) models, an exponential smoothing model, and a segmented trend forecasting model | |
| **VTSanghani-ExogModel** (2022-23) | AI | Deep learning model that uses various exogenous datasets | Air Quality Index, and HHS confirmed COVID-19 hospitalizations at the state level and aggregated to weeks. |

*Data used in addition to the HHS COVID-19 Reported Patient Impact and Hospital Capacity by State Timeseries – confirmed influenza hospitalization admissions

Table S2: Types of models submitted to FluSight but did not meet inclusion criteria.

| Model Name | Type | Methods | Additional data used* | Repo_url |
|---|---|---|---|---|
| **CADPH-FluCAT_Ensemble** (2022-23) | TS Ensemble | This model is an ensemble of several time series forecasting and predictive models targeting influenza hospital admits based on historical clinical lab and public health lab flu surveillance data. | California Department of Health Care Access and Information | https://calcat.covid19.ca.gov/cacovidmodels/ |
| **CEID-Walk** (2022-23) | Statistical | Random walk model without drift. The last few observations of a target time series are used to estimate the variance in step size of the random walk. | | |
| **ISU_NiemiLab-Flu** (2022-23) | Statistical | By state ARIMA | | |
| **JHUAPL-Gecko** (2021-22) | Time Series | SARIMA time series model (1,1,0)x(1,1,0,7) with anomaly detector applied to confirmed hospital admissions since 09/01/2020. | | https://gitlab.jhuapl.edu/panagmj1/gecko-sarima |
| **LosAlamos_NAU-Cmodel_Flu** (2022-23) | Mechanistic Statistical | Seasonal compartmental model to perform Bayesian analysis on hospitalization data available for each state. | | https://github.com/computationalUncertaintyLab/2022Flu_LucompUncertLab |
| **LUcompUncertLab-ensemble_rclp** (2022-23) | Statistical Ensemble | A model that combines component models and recalibrates using a mixture of beta distributions | Models from LU | https://github.com/computationalUncertaintyLab/ |
| **LUcompUncertLab-experthuman** (2022-23) | Human Judgement | A model built by expert judgement predictions. | Expert judgement prob densities | https://github.com/computationalUncertaintyLab/2022Flu_LUcompUncertLab |
| **LUcompUncertLab-hier_mech_model** (2022-23) | Hierarchical Mechanistic | We fit a SIRHD compartmental model where S0 is shared between seasons | Weekly num of ILI from ILInet | https://github.com/computationalUncertaintyLab/ |
| **LucompUncertLab-humanjudgement** (2021-22, 2022-23) | Human judgement | A model built by interpolating Human judgement predictions and projecting them from six locations to all 54. | Human judgement probability densities from Metaculus | https://github.com/lanl/SeasonalFlu-2022-Predictions |
| **LUcompUncertLab-HWAR2** (2022-23) | Statistical | We detrend with Holt Winters and then fit the residuals with a Bayesian AR(2) process | | https://github.com/computationalUncertaintyLab/ |
| **LUcompUncertLab-KalmanFilter** (2022-23) | Statistical | A model that supposes there is a ltent AR process that generates observations. | | https://github.com/computationalUncertaintyLab/ |
| **LUcompUncertLab-stacked_ili** (2022-23) | | A model that incorporates ILI data | | https://github.com/computationalUncertaintyLab/ |

| | | | | |
|---|---|---|---|---|
| **NIH-Flu_ARIMA** (2022-23) | Statistical | Seasonal ARIMA model with exogenous covariates for cumulative vaccination coverage, seasonal vaccine effectiveness, weekly A/H3N2 circulation, and influenza transmissibility. | CDC FluSurv-NET hospitalization rates, syndromic and virologic surveillance data from CDC FluView, vaccination coverage estimates from the National Center for Immunization and Respiratory Diseases, and seasonal vaccine effectiveness estimates from published observational studies and the CDC | https://github.com/midas-network/flu-scenario-modeling-hub/tree/main/data-processed/NIH-Flu_TS |
| **UGuelph-FluPLUG** (2022-23) | TS Statistical | Applies piecewise linear segmentation to time series of log(incidence) informed also by data on past seasons. | | https://sites.google.com/site/mgcojocarumath/networks-and-dynamics-lab |
| **Umass-ARIMA** (2021-22) | TS regression Statistcal | Autoregressive integrated moving average of time-series model | MA DPH Syndromic Surveillance (SyS) for 2017 to 2021 | https://github.com/reichlab/flu-hosp-models-2021-22 |
| **UMass-gbq** (2022-23) | Statistical | Gradient boosting for the median, followed by conformal prediction to get quantiles. | | https://github.com/reichlab/covid-hosp-models |
| **UNC_IDD-InfluPaint** (2022-23) | Statistical | We build a generative denoising diffusion probabilistic model of Influenza dynamics that generates synthetic epidemic trajectories. We condition these trajectories on ground-truth data using an inpainting algorithm to produce our forecasts. | | https://github.com/jcblemai/inpainting-idforecasts |
| **VTSanghani-ExogModel** (2021-22) | AI | Deep learning model that uses various exogenous datasets | Air Quality Index, and HHS confirmed COVID-19 hospitalizations at the state level and aggregated to weeks. | |
| **VTSanghani-Transformer** (2022-23) | TS Statistical | A model based on the Temporal Fusion Transformer architecture. | | https://sanghani.cs.vt.edu/ |

Figure S1: Distribution of relative WIS across models for all subnational forecast jurisdictions. The horizontal blue line is at one, where forecast performance is the same as the Flusight-baseline (see Figure 3).

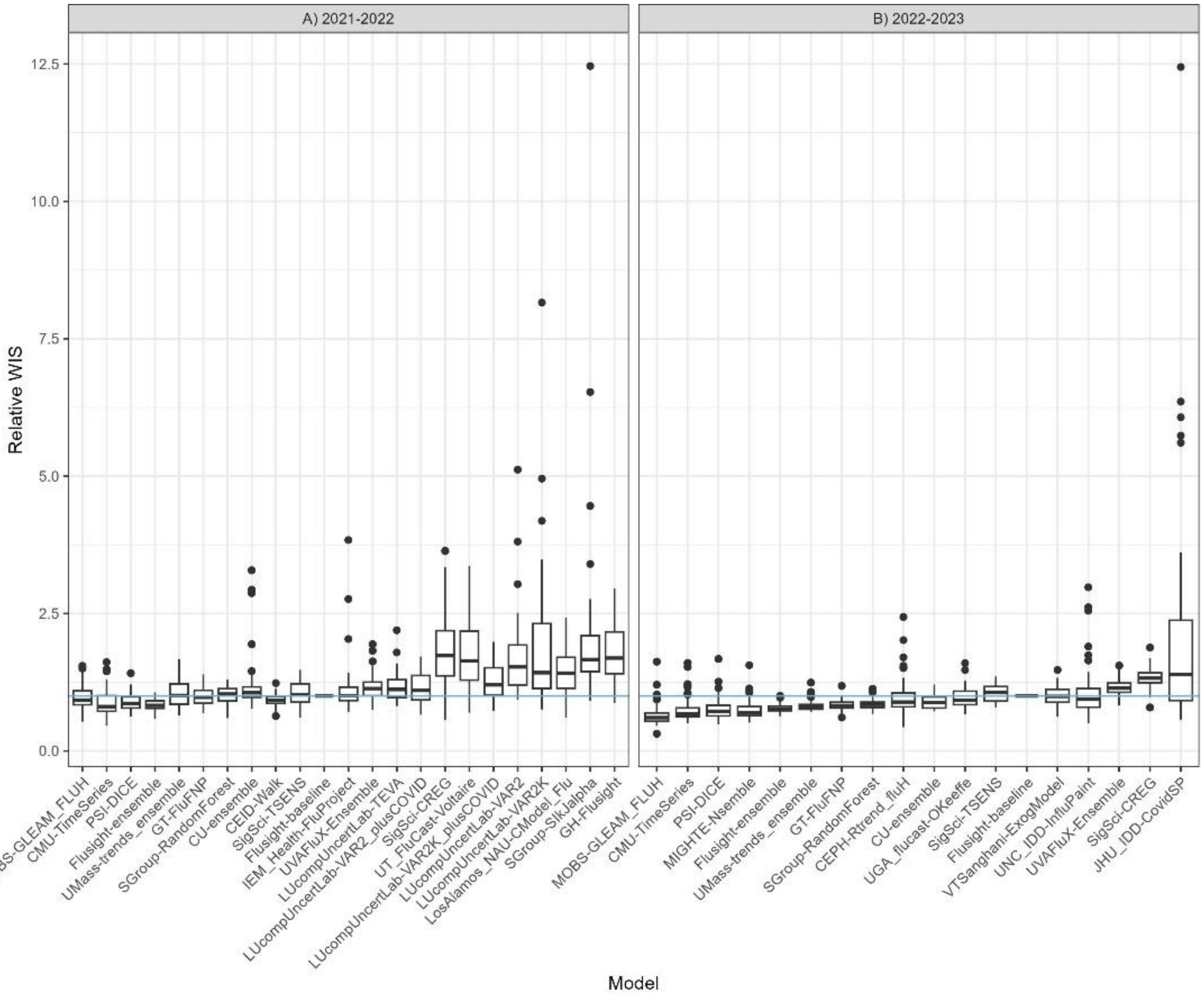

### Supplemental Analysis 1: Backfill/Revision of hospital admission data

#### Methods

Differences between the finalized data used in this evaluation (reported as of September 12, 2022, for the 2021-22 season, and June 13, 2023, for the 2022-23 season) and the originally reported values were analyzed by comparing differences in the weekly incident rate by both the relative change and absolute change. Original reported confirmed influenza hospital admissions were downloaded from the COVID-19 Reported Patient Impact and Hospital Capacity by State Timeseries Archive Repository [7] and aggregated by week for each state and territory. Reported values of weekly hospitalizations were compared each week for the entire U.S. from February 7, 2022, to July 16, 2022, for the 2021-22 season, and October 17, 2022, to June 13, 2023 for the 2022-23 season. This was done to capture updates spanning the start of the mandatory influenza reporting period through the weeks needed to evaluate the last four-week ahead forecasts. For each jurisdiction, we calculated the percent difference and numeric difference between the number of hospitalizations between the initial week of publication and the number of hospitalizations when the data were considered final for these analyses. During the 2022-23 season, it became evident that the frequency and magnitude of backfilled counts was dependent on day of the week. Revisions were more likely to occur on Tuesdays rather than on Mondays. Due to this day of the week revision effect, weekly observed data used in this analysis reflect the days that data were pulled during the season; prior to January 16, data were downloaded each Monday, and beginning January 16, data were downloaded each Tuesday.

#### Results

To assess potential impacts of changes in reported values on forecast performance, we compared reported incident hospitalizations [8] at the time they would have been used for forecasts to the values used in evaluating forecasts (Figures S2-S4). Most data updates occurred within one month of when the data were initially published, and 12 out of the 18 weeks that were analyzed did not have updates two weeks after initial publication for 2021-22. Prior to the week ending December 19, most data updates occurred within one month of when the data were initially published. Subsequently, data were updated more frequently over a longer period. For both seasons, across all subnational forecast jurisdictions, a majority (93.96% in 2021-22 and 83.48% in 2022-23) of final values changed by less than ten hospitalizations or did not change compared to corresponding initial reports. In 2021-22, a higher percent of updates had final values lower than initial reports, 37.91%, compared to 30.37% in 2022-23. Conversely, 2022-23 had a higher percent of updates with no change (38.69%) than the 2021-22 season (31.68%). Both seasons had similar percent of updates with final values higher than initial reports (30.94% and 30.4%, for 2022-23 and 2021-22 respectively). For the U.S., the median percent and numeric difference across all forecast weeks were lower in 2021-22, with a median percent difference of -2.1% (range: -4.75% to 0.6%), and the median numeric difference of -39 (range: -157, 19). For 2022-23, the median percent difference across all forecast weeks was 0.46% (range: -7.93% to 6.55%), and the median numeric difference was 14.5 (range: -389, 1,220).

Jurisdiction-specific median percent differences had a larger range in 2021-22 from -14.29% in Rhode Island to 7.14% in Oregon whereas percent differences ranged from -4.53% in Indiana to 4.39% in Illinois for 2022-23. Across states, DC, and Puerto Rico, and for both seasons, the median numeric difference between initial report and final report was 0. Figure S2- Figure S4 for additional information on the distribution of differences between final reported values used for evaluation and the initial reported values.

Figure S2: Data updates analysis: initial epicurve (points available at time of forecast in blue) and final epicurve reported from HHS-Protect as of the final evaluation date of September 12, 2022 and June 12, 2023 for each season, respectively (in red).

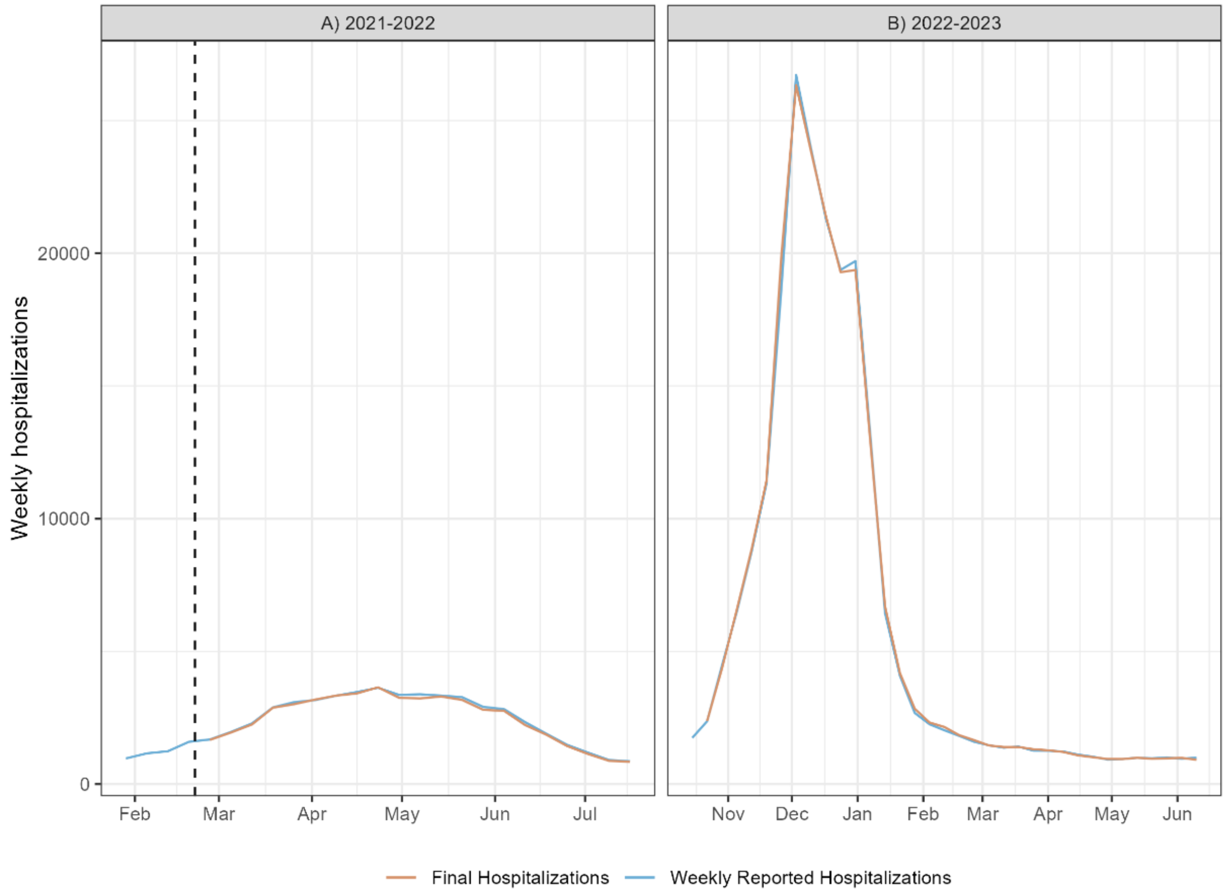

Figure S3: state-specific distributions of absolute and relative changes in HHS-Protect reported hospitalizations as of the initial report date and the final evaluation date (September 12, 2022).

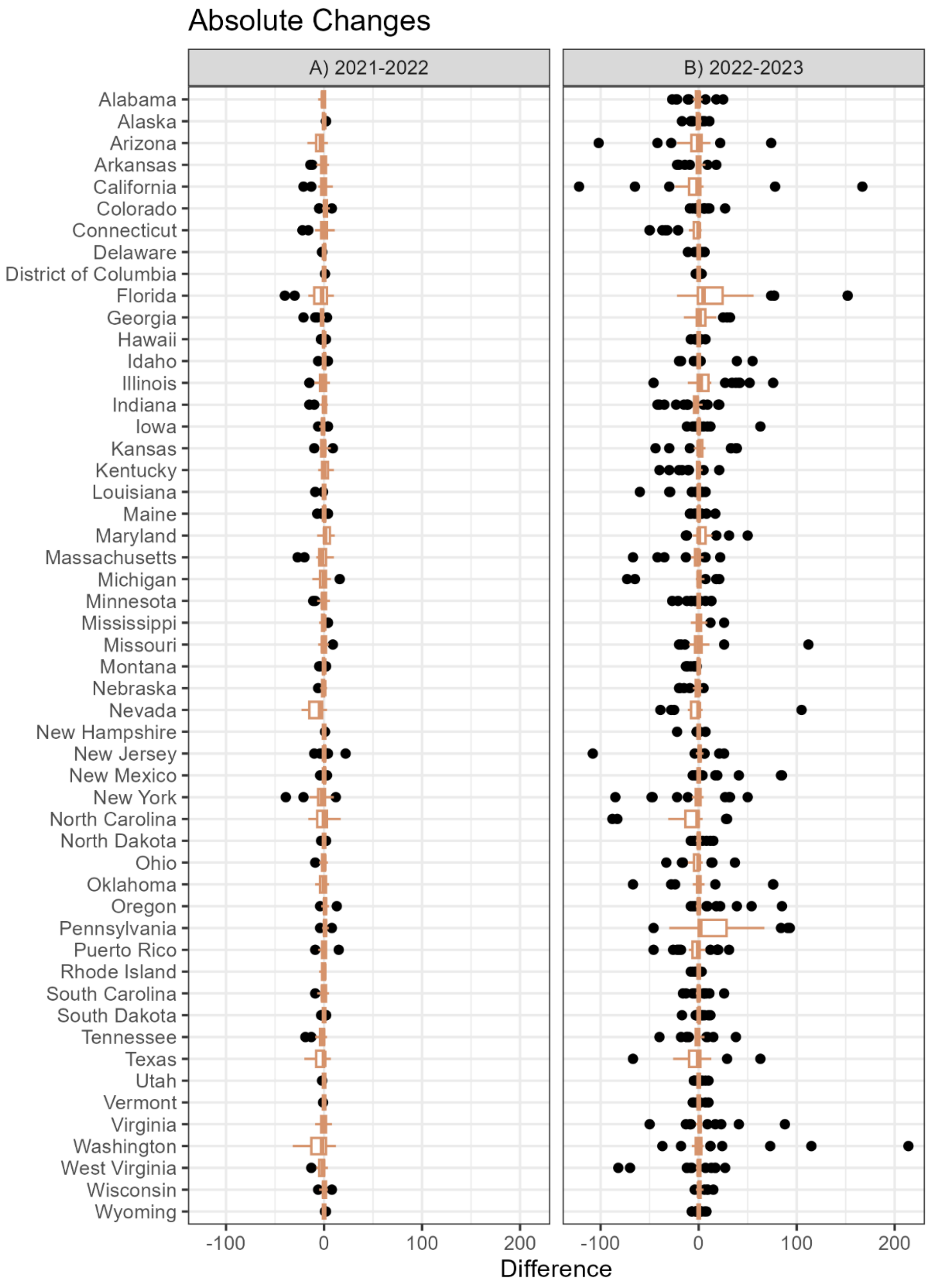

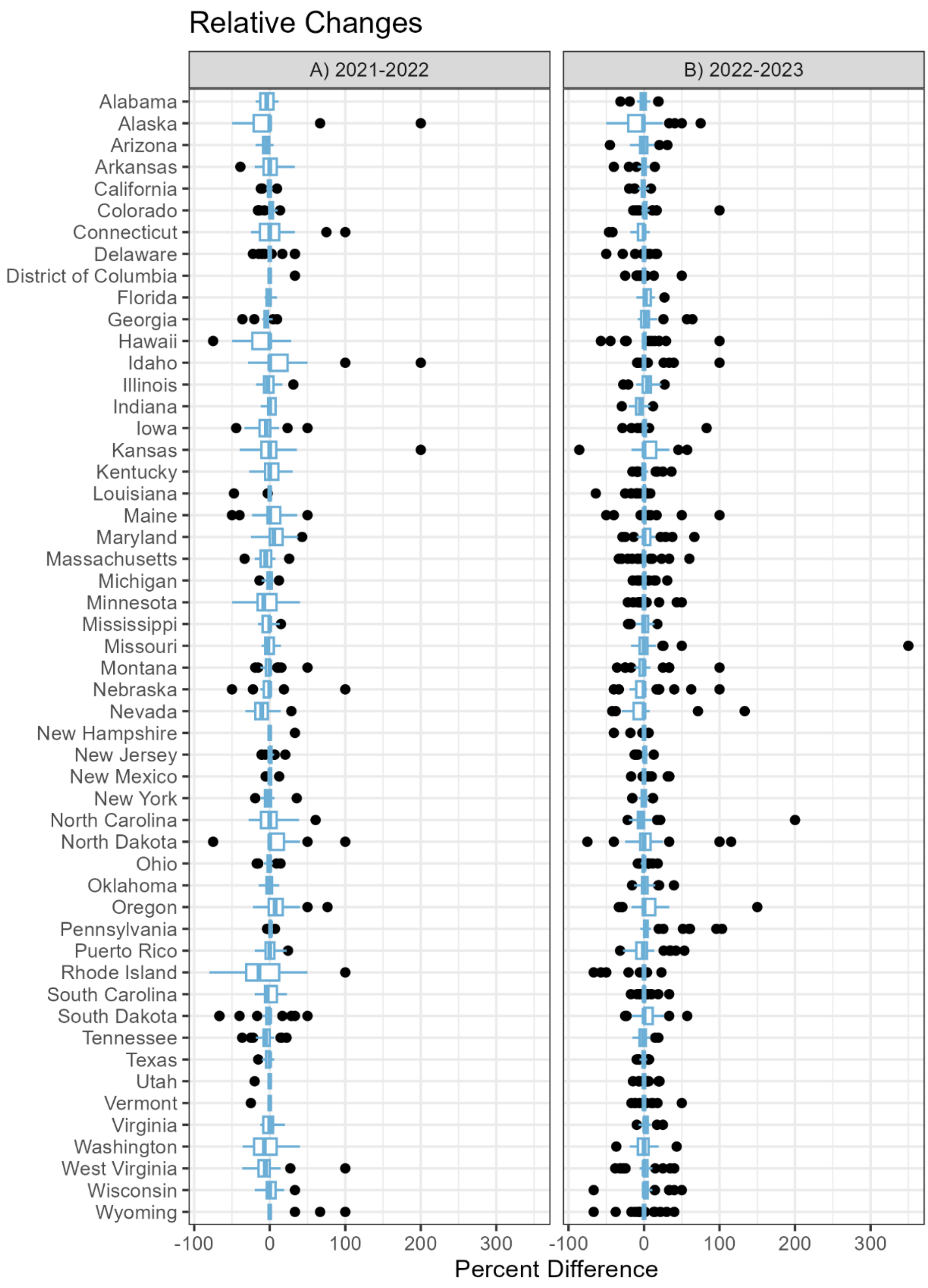

Relative Changes
A) 2021-2022
B) 2022-2023
Alabama
Alaska
Arizona
Arkansas
California
Colorado
Connecticut
Delaware
District of Columbia
Florida
Georgia
Hawaii
Idaho
Illinois
Indiana
Iowa
Kansas
Kentucky
Louisiana
Maine
Maryland
Massachusetts
Michigan
Minnesota
Mississippi
Missouri
Montana
Nebraska
Nevada
New Hampshire
New Jersey
New Mexico
New York
North Carolina
North Dakota
Ohio
Oklahoma
Oregon
Pennsylvania
Puerto Rico
Rhode Island
South Carolina
South Dakota
Tennessee
Texas
Utah
Vermont
Virginia
Washington
West Virginia
Wisconsin
Wyoming
-100
0
100
200
300
Percent Difference

Figure S4: Data updates analysis: U.S. weekly reported hospitalizations from from the HHS-Protect system [8] with the initial date each weeks' data were pulled (vertical axis) and reported values as of subsequent dates (horizontal axis). Coloring for the number of hospitalizations was normalized on a common log scale.

**a)** 2021-22 season

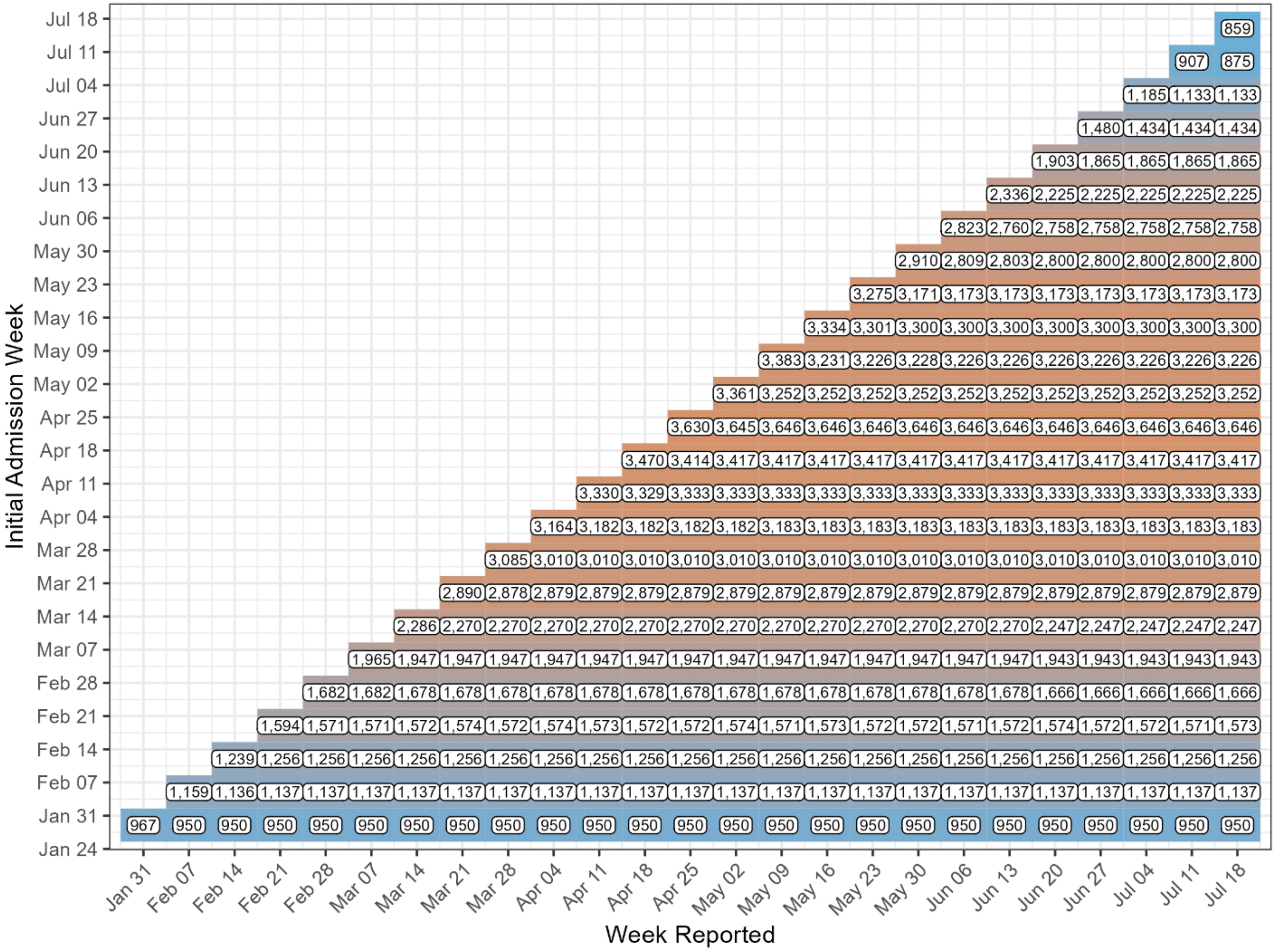

**b)** 2022-23 season

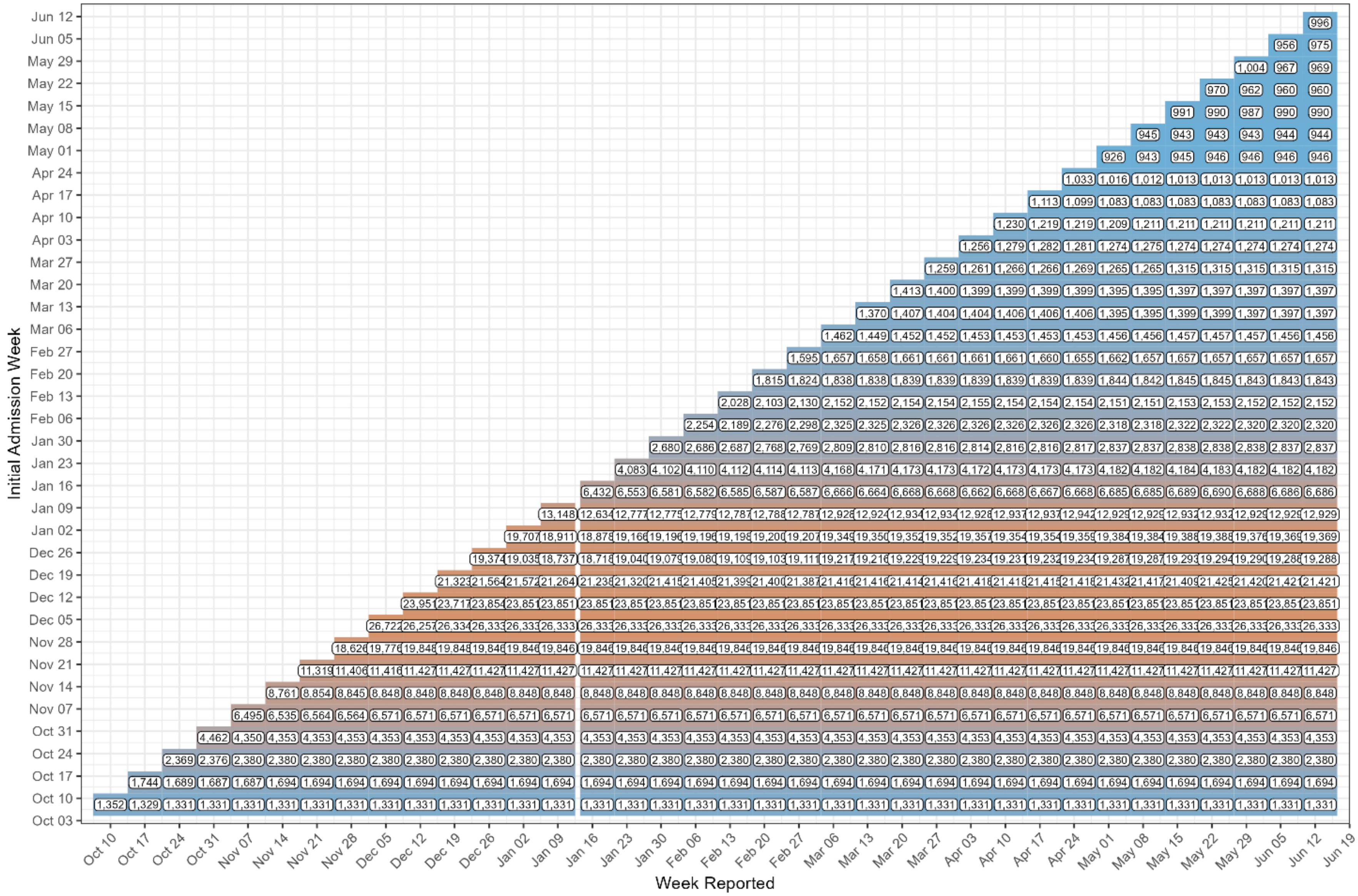

### Supplemental Analysis 2: Performance of log-transformed forecasts

Using the same methods described in the methods section, we analyzed performance based on log-transformed hospitalization counts. In terms of absolute and relative WIS, teams that outperformed the FluSight Ensemble in the regular analysis also outperformed the ensemble in the log-transformed analysis. For each season, the same five teams performed best in both analyses. In 2021-22, 8 models outperformed the baseline, compared to 6 when not transformed. In 2022-23, 11 models outperformed the baseline, compared to 12 when not transformed (Table S4).

In terms of standardized rank, the FluSight Ensemble outperformed the other models for both seasons. Other models ranked similarly between analyses (Figure S5).

Coverage was similar when using log-transformed hospitalization counts.

Table S3

| Model | Absolute WIS | Relative WIS | MAE | 50% Coverage (%) | 95% Coverage (%) | % of Forecasts Submitted |
|---|---|---|---|---|---|---|
| **2021-2022** | | | | | | |
| CMU-TimeSeries | 0.31 | 0.78 | 0.47 | 47 | 90 | 100 |
| Flusight-ensemble | 0.33 | 0.83 | 0.50 | 48 | 86 | 100 |
| PSI-DICE | 0.33 | 0.84 | 0.50 | 43 | 82 | 100 |
| UMass-trends_ensemble | 0.36 | 0.91 | 0.53 | 71 | 97 | 100 |
| SGroup-RandomForest | 0.38 | 0.97 | 0.59 | 47 | 95 | 100 |
| CU-ensemble | 0.39 | 0.98 | 0.57 | 44 | 77 | 100 |
| GT-FluFNP | 0.38 | 0.98 | 0.54 | 39 | 69 | 96 |
| CEID-Walk | 0.39 | 0.99 | 0.55 | 52 | 82 | 89 |
| Flusight-baseline | 0.40 | 1.00 | 0.56 | 49 | 83 | 100 |
| SigSci-TSENS | 0.40 | 1.01 | 0.55 | 38 | 72 | 96 |
| IEM_Health-FluProject | 0.40 | 1.02 | 0.58 | 50 | 85 | 100 |
| LUcompUncertLab-TEVA | 0.41 | 1.05 | 0.59 | 54 | 86 | 89 |
| LUcompUncertLab-VAR2_plusCOVID | 0.42 | 1.08 | 0.60 | 42 | 74 | 94 |
| MOBS-GLEAM_FLUH | 0.42 | 1.08 | 0.59 | 32 | 63 | 91 |
| UT_FluCast-Voltaire | 0.45 | 1.14 | 0.71 | 50 | 91 | 99 |
| UVAFluX-Ensemble | 0.45 | 1.14 | 0.57 | 38 | 64 | 99 |
| LUcompUncertLab-VAR2K_plusCOVID | 0.47 | 1.20 | 0.68 | 42 | 74 | 89 |
| SGroup-SIkJalpha | 0.49 | 1.24 | 0.63 | 18 | 46 | 100 |
| LUcompUncertLab-VAR2 | 0.53 | 1.35 | 0.77 | 39 | 72 | 94 |
| LUcompUncertLab-VAR2K | 0.61 | 1.56 | 0.93 | 42 | 83 | 89 |

| Model | Absolute WIS | Relative WIS | MAE | 50% Coverage (%) | 95% Coverage (%) | % of Forecasts Submitted |
|---|---|---|---|---|---|---|
| LosAlamos_NAU-CModel_Flu | 0.63 | 1.60 | 0.90 | 26 | 59 | 100 |
| GH-Flusight | 0.74 | 1.88 | 0.79 | 6 | 13 | 94 |
| SigSci-CREG | 0.80 | 2.03 | 1.04 | 19 | 44 | 89 |
| **2022-2023** | | | | | | |
| MOBS-GLEAM_FLUH | 0.37 | 0.65 | 0.54 | 38 | 75 | 93 |
| CMU-TimeSeries | 0.41 | 0.70 | 0.62 | 46 | 84 | 94 |
| MIGHTE-Nsemble | 0.41 | 0.70 | 0.58 | 48 | 79 | 96 |
| PSI-DICE | 0.42 | 0.71 | 0.60 | 45 | 77 | 100 |
| Flusight-ensemble | 0.44 | 0.74 | 0.62 | 51 | 77 | 100 |
| JHU_IDD-CovidSP | 0.49 | 0.82 | 0.69 | 47 | 78 | 83 |
| UMass-trends_ensemble | 0.49 | 0.83 | 0.69 | 60 | 87 | 100 |
| CU-ensemble | 0.51 | 0.85 | 0.69 | 45 | 66 | 85 |
| SGroup-RandomForest | 0.52 | 0.88 | 0.75 | 48 | 81 | 96 |
| GT-FluFNP | 0.53 | 0.89 | 0.66 | 52 | 72 | 95 |
| UNC_IDD-InfluPaint | 0.52 | 0.94 | 0.74 | 36 | 71 | 75 |
| Flusight-baseline | 0.59 | 1.00 | 0.73 | 41 | 68 | 100 |
| UGA_flucast-OKeeffe | 0.61 | 1.02 | 0.82 | 41 | 67 | 94 |
| UVAFluX-Ensemble | 0.61 | 1.03 | 0.75 | 20 | 39 | 98 |
| CEPH-Rtrend_fluH | 0.58 | 1.05 | 0.89 | 38 | 74 | 88 |
| VTSanghani-ExogModel | 0.63 | 1.05 | 0.83 | 27 | 58 | 81 |
| SigSci-TSENS | 0.66 | 1.09 | 0.88 | 50 | 68 | 93 |
| SigSci-CREG | 0.68 | 1.16 | 0.83 | 32 | 54 | 90 |
| ISU_NiemiLab-Flu | 0.71 | 1.25 | 0.93 | 29 | 51 | 77 |

The Absolute WIS column refers to the Weighted Interval Score for each model across all fifty states, D.C., and Puerto Rico for log-transformed forecast targets. The Relative WIS compares the WIS value of each model to the Flusight-baseline model for log-transformed forecast targets. All models with a relative WIS score less than one outperformed the baseline model when evaluated solely upon WIS. 95% and 50% coverage values are provided for the percent of times that reported weekly incidence values were within the 95% or 50% prediction intervals respectively, across all the forecast targets submitted by each team. The percent of forecasts submitted is determined by the number of forecast targets submitted by each team out of all possible forecast targets occurring within the duration of the evaluation period.

Table S4

| Model | Relative WIS | % WIS Below Baseline | 1 Wk Coverage | 2 Wk Coverage | 3 Wk Coverage | 4 Wk Coverage | % Cov abv 90 (1 Wk) | % Cov abv 90 (2 Wk) | % Cov abv 90 (3 Wk) | % Cov abv 90 (4 Wk) |
|---|---|---|---|---|---|---|---|---|---|---|
| **2021-2022** | | | | | | | | | | |
| CMU-TimeSeries | 0.78 | 88.46 | 90.17 | 91.45 | 90.60 | 86.54 | 50.00 | 72.22 | 61.11 | 27.78 |
| Flusight-ensemble | 0.83 | 96.15 | 89.32 | 86.11 | 85.15 | 83.33 | 55.56 | 33.33 | 27.78 | 38.89 |
| PSI-DICE | 0.84 | 86.54 | 88.89 | 83.87 | 78.31 | 76.50 | 38.89 | 27.78 | 5.56 | 0.00 |
| UMass-trends_ensemble | 0.91 | 63.46 | 96.15 | 97.65 | 96.90 | 96.15 | 100.00 | 100.00 | 100.00 | 100.00 |
| SGroup-RandomForest | 0.97 | 65.38 | 95.41 | 94.87 | 94.66 | 94.12 | 88.89 | 88.89 | 83.33 | 88.89 |
| CU-ensemble | 0.98 | 59.62 | 79.59 | 80.66 | 76.50 | 71.90 | 16.67 | 11.11 | 0.00 | 0.00 |
| GT-FluFNP | 0.98 | 60.00 | 70.11 | 68.67 | 68.22 | 70.11 | 5.56 | 16.67 | 16.67 | 22.22 |
| CEID-Walk | 0.99 | 59.62 | 82.09 | 83.77 | 81.01 | 81.85 | 37.50 | 37.50 | 31.25 | 37.50 |
| Flusight-baseline | 1.00 | 0.00 | 82.26 | 84.19 | 82.48 | 81.62 | 27.78 | 22.22 | 22.22 | 22.22 |
| SigSci-TSENS | 1.01 | 52.00 | 74.11 | 73.44 | 70.54 | 69.20 | 11.11 | 5.56 | 5.56 | 5.56 |
| IEM_Health-FluProject | 1.02 | 50.00 | 91.45 | 86.54 | 82.59 | 78.21 | 72.22 | 38.89 | 22.22 | 22.22 |
| LUcompUncertLab-TEVA | 1.05 | 38.46 | 84.86 | 85.58 | 86.06 | 86.18 | 25.00 | 18.75 | 25.00 | 31.25 |
| LUcompUncertLab-VAR2_plusCOVID | 1.08 | 50.00 | 76.70 | 74.77 | 73.30 | 70.14 | 17.65 | 5.88 | 5.88 | 5.88 |
| MOBS-GLEAM_FLUH | 1.08 | 36.00 | 71.11 | 65.80 | 59.79 | 56.49 | 0.00 | 0.00 | 0.00 | 0.00 |
| UT_FluCast-Voltaire | 1.14 | 30.77 | 94.73 | 90.96 | 89.13 | 90.42 | 83.33 | 72.22 | 55.56 | 61.11 |
| UVAFluX-Ensemble | 1.14 | 17.31 | 66.05 | 65.51 | 62.58 | 60.95 | 11.11 | 0.00 | 0.00 | 0.00 |
| LUcompUncertLab-VAR2K_plusCOVID | 1.20 | 28.85 | 75.72 | 75.24 | 74.04 | 72.72 | 6.25 | 0.00 | 0.00 | 0.00 |
| SGroup-SIkJalpha | 1.24 | 7.69 | 40.28 | 45.73 | 48.08 | 48.29 | 0.00 | 0.00 | 0.00 | 0.00 |
| LUcompUncertLab-VAR2 | 1.35 | 5.77 | 73.87 | 72.29 | 72.17 | 70.81 | 11.76 | 5.88 | 11.76 | 11.76 |
| LUcompUncertLab-VAR2K | 1.56 | 7.69 | 81.97 | 81.49 | 83.05 | 85.46 | 6.25 | 18.75 | 25.00 | 37.50 |
| LosAlamos_NAU-CModel_Flu | 1.60 | 13.46 | 65.28 | 59.29 | 56.52 | 54.06 | 5.56 | 0.00 | 0.00 | 0.00 |
| GH-Flusight | 1.88 | 3.85 | 18.33 | 12.90 | 11.99 | 10.63 | 0.00 | 0.00 | 0.00 | 0.00 |
| SigSci-CREG | 2.03 | 8.00 | 46.87 | 43.98 | 43.86 | 43.13 | 0.00 | 0.00 | 0.00 | 0.00 |
| **2022-2023** | | | | | | | | | | |
| MOBS-GLEAM_FLUH | 0.65 | 100.00 | 78.42 | 74.60 | 73.41 | 75.32 | 30.77 | 19.23 | 15.38 | 25.81 |
| CMU-TimeSeries | 0.70 | 94.23 | 84.13 | 84.84 | 84.76 | 84.13 | 50.00 | 57.69 | 65.38 | 60.00 |

| Model | Relative WIS | % WIS Below Baseline | 1 Wk Coverage | 2 Wk Coverage | 3 Wk Coverage | 4 Wk Coverage | % Cov abv 90 (1 Wk) | % Cov abv 90 (2 Wk) | % Cov abv 90 (3 Wk) | % Cov abv 90 (4 Wk) |
|---|---|---|---|---|---|---|---|---|---|---|
| MIGHTE-Nsemble | 0.70 | 100.00 | 83.65 | 81.19 | 78.10 | 72.22 | 56.00 | 52.00 | 60.00 | 58.06 |
| PSI-DICE | 0.71 | 98.08 | 86.09 | 78.18 | 73.22 | 71.60 | 57.69 | 61.54 | 57.69 | 70.97 |
| Flusight-ensemble | 0.74 | 100.00 | 83.14 | 78.18 | 74.70 | 73.59 | 57.69 | 61.54 | 57.69 | 61.29 |
| JHU_IDD-CovidSP | 0.82 | 72.55 | 84.86 | 79.03 | 75.27 | 72.04 | 59.09 | 54.55 | 50.00 | 70.97 |
| UMass-trends_ensemble | 0.83 | 92.31 | 90.01 | 88.24 | 85.21 | 83.65 | 73.08 | 69.23 | 65.38 | 48.00 |
| CU-ensemble | 0.85 | 92.31 | 67.83 | 67.05 | 65.12 | 62.76 | 36.36 | 45.45 | 45.45 | 53.85 |
| SGroup-RandomForest | 0.88 | 92.31 | 88.08 | 81.38 | 78.23 | 76.46 | 68.00 | 64.00 | 64.00 | 66.67 |
| GT-FluFNP | 0.89 | 86.00 | 73.24 | 69.58 | 72.07 | 74.65 | 50.00 | 50.00 | 50.00 | 65.52 |
| UNC_IDD-InfluPaint | 0.94 | 70.59 | 71.93 | 70.56 | 71.05 | 71.84 | 55.00 | 40.00 | 55.00 | 56.00 |
| Flusight-baseline | 1.00 | 0.00 | 74.63 | 69.30 | 65.83 | 64.05 | 50.00 | 50.00 | 50.00 | 58.06 |
| UGA_flucast-OKeeffe | 1.02 | 52.94 | 76.31 | 67.69 | 62.75 | 60.39 | 40.00 | 36.00 | 28.00 | 0.00 |
| UVAFluX-Ensemble | 1.03 | 37.25 | 40.87 | 41.25 | 37.03 | 37.03 | 0.00 | 0.00 | 0.00 | 46.43 |
| CEPH-Rtrend_fluH | 1.05 | 50.00 | 70.99 | 75.92 | 74.83 | 73.24 | 34.78 | 39.13 | 47.83 | 4.00 |
| VTSanghani-ExogModel | 1.05 | 40.38 | 63.00 | 58.15 | 55.31 | 55.13 | 0.00 | 0.00 | 0.00 | 40.00 |
| SigSci-TSENS | 1.09 | 12.00 | 71.60 | 68.97 | 67.54 | 65.63 | 46.15 | 46.15 | 46.15 | 45.16 |
| SigSci-CREG | 1.16 | 14.00 | 61.83 | 54.74 | 50.70 | 48.15 | 38.46 | 38.46 | 34.62 | 58.06 |
| ISU_NiemiLab-Flu | 1.25 | 3.85 | 54.81 | 55.19 | 49.42 | 46.35 | 20.00 | 25.00 | 10.00 | |

The % WIS Below Baseline column shows the percent of WIS values for each model below the corresponding FluSight-Baseline WIS. The '% Cov abv 90' columns show the percent of weekly 95% coverage values that are greater than or equal to 90% for each model by horizon.

Figure S5: Standardized rank of weighted interval score (WIS) for log-transformed hospital admission forecasts over all forecast jurisdictions and horizons (1- to 4-week ahead), for the FluSight ensemble and each team submitting at least 75% of the forecast targets (see Table 1 for qualifying teams and season metrics).

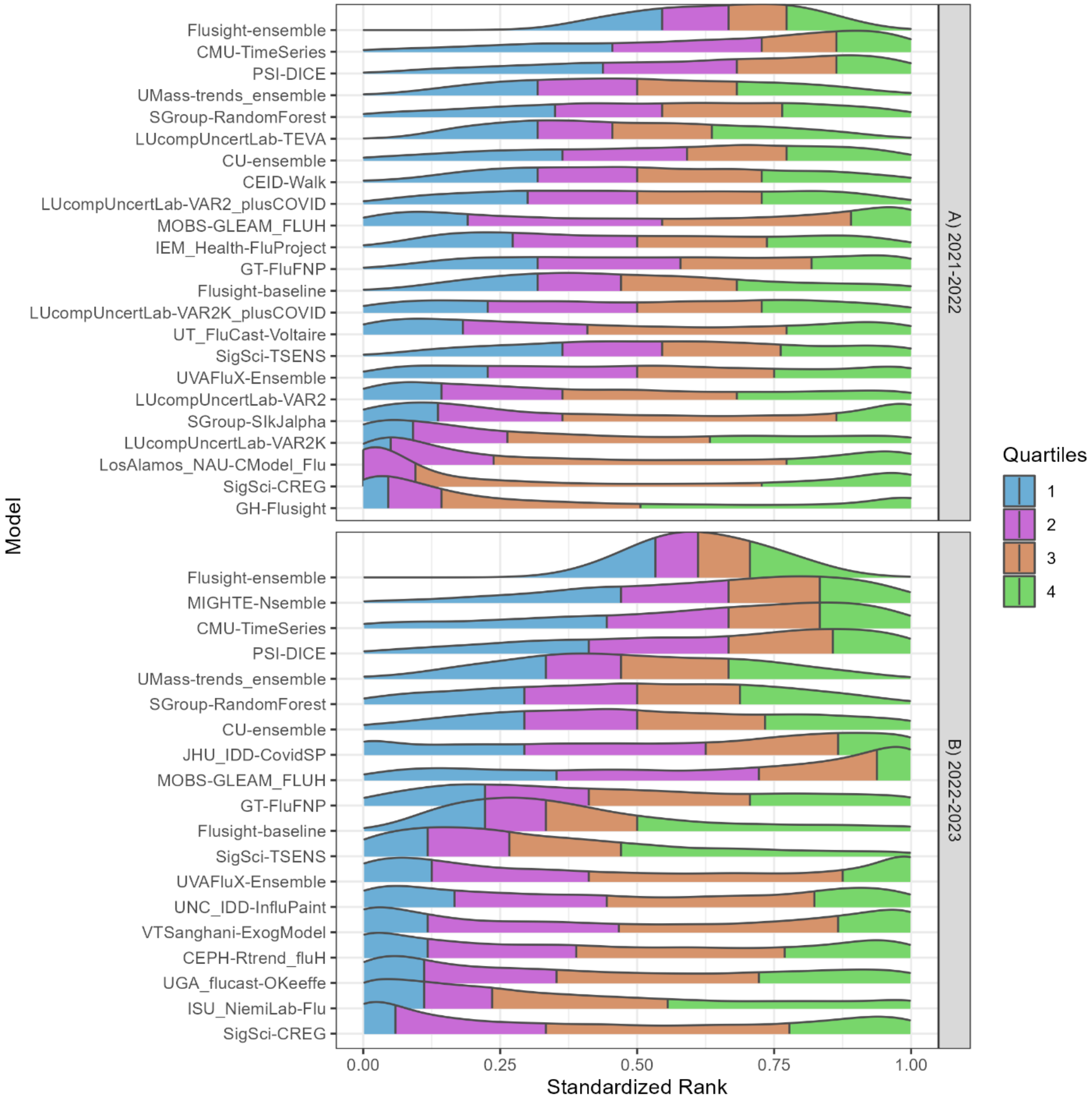

Figure S6: Time series of absolute WIS of log-transformed forecasts. Note that the forecast evaluation period translates to 1-week ahead forecast target end dates from February 26 to June 25, 2022, and October 22, 2022, to May 20, 2023, and 4-week ahead forecast target end dates from March 19 to July 16, 2022, and November 5, 2022, to June 10, 2023. Weekly results for the FluSight baseline and ensemble models are shown in red and blue respectively. Results for individual contributing models are shown in light gray.

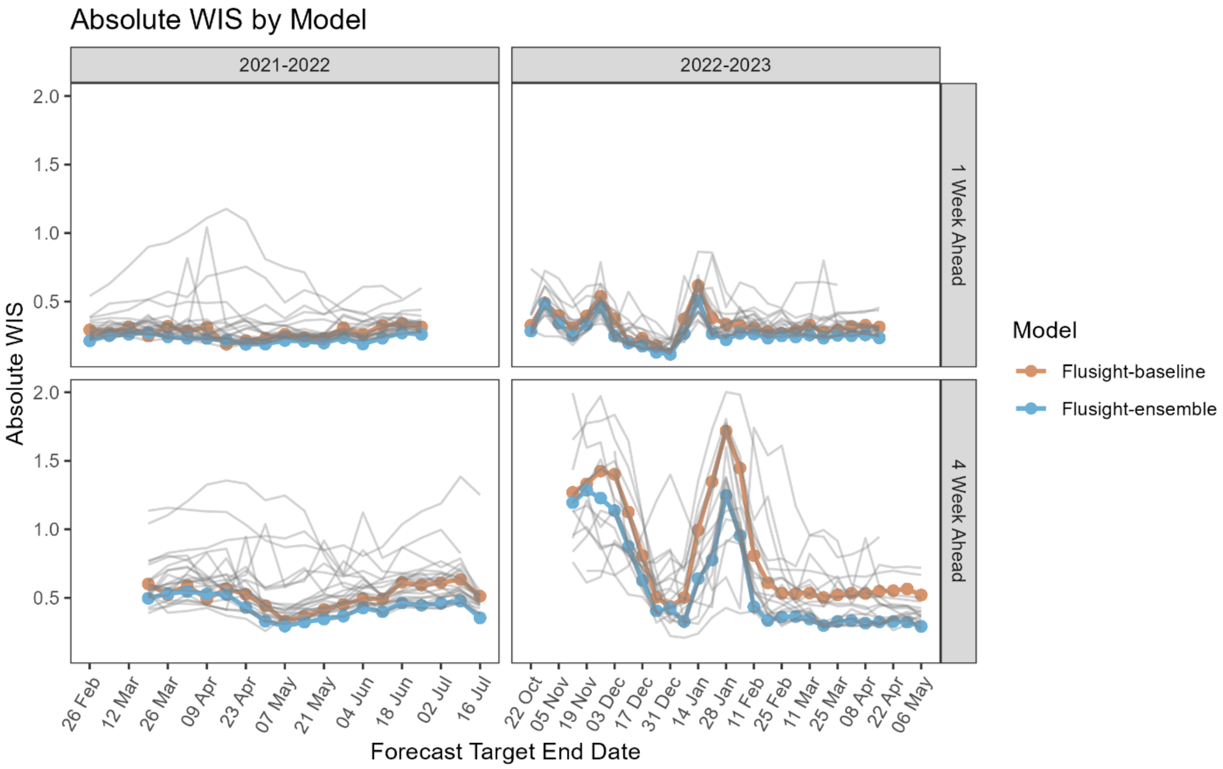

Figure S7: State-level WIS values for each team relative to the FluSight baseline model using log-transformed hospitalization counts. The range of Relative WIS values below 1, in blue, indicate better performance than the FluSight baseline (white). Relative WIS values above 1, in red, indicate poor performance relative to the FluSight baseline. Teams are ordered on the horizontal axis from lowest to highest Relative WIS values for each season.

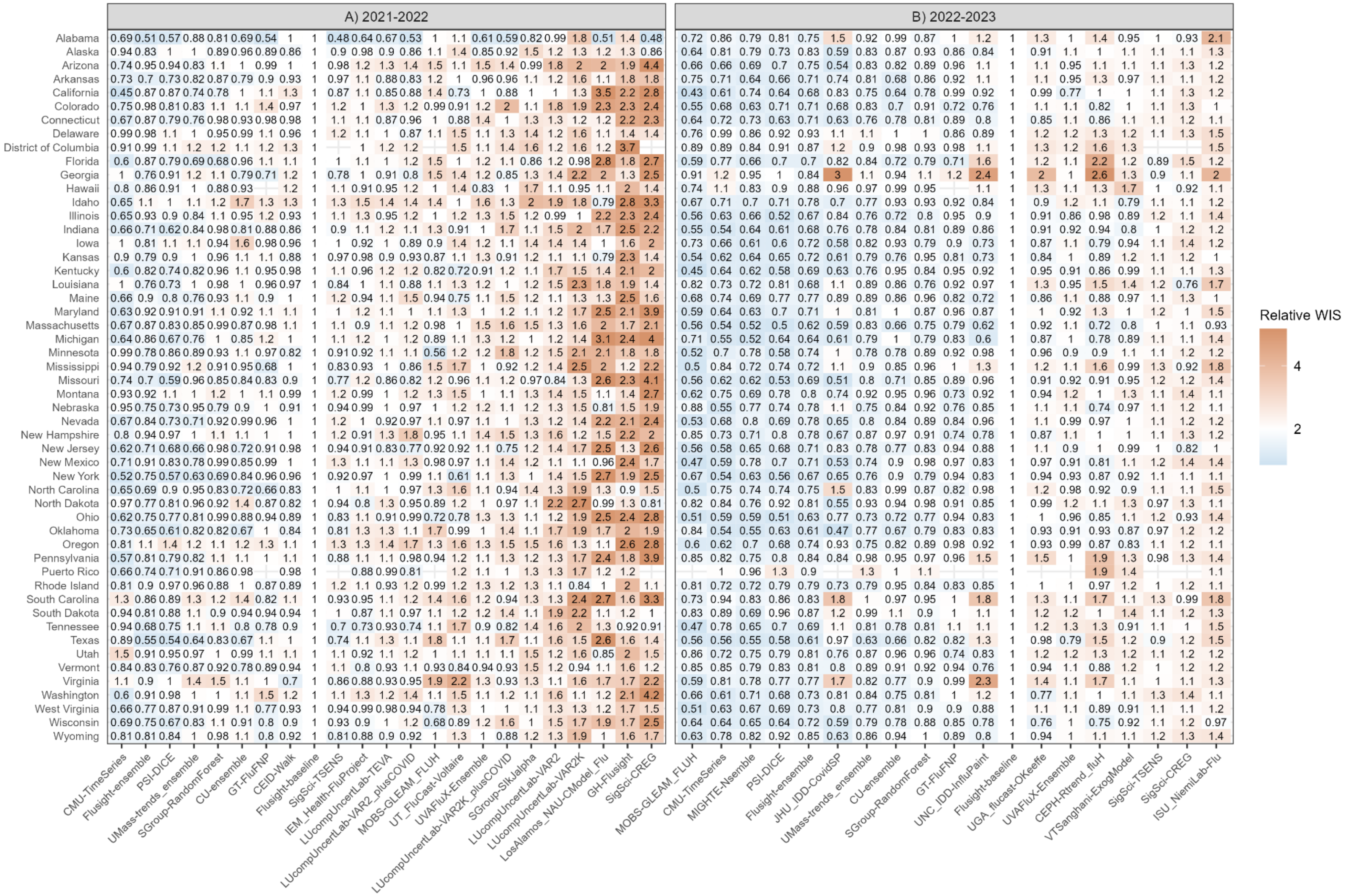

### Supplement Analysis 3: Performance of national forecasts

When considering just national forecast targets, two additional teams met the eligibility criteria for inclusion in the end of season analysis. These models, as well as others submitting for a subset of jurisdictions, were included when generating ensembles during the season.

Table S5: Performance metrics for teams submitting at least 75% of weekly national FluSight targets.

| Model | Absolute WIS | Relative WIS | MAE | 50% Coverage (%) | 95% Coverage (%) | % of Forecasts Submitted |
|---|---|---|---|---|---|---|
| **2021-22** | | | | | | |
| PSI-DICE | 296.70 | 0.71 | 421.38 | 56 | 89 | 100 |
| UMass-trends_ensemble | 301.67 | 0.72 | 485.97 | 46 | 85 | 100 |
| CMU-TimeSeries | 316.02 | 0.75 | 355.28 | 88 | 100 | 100 |
| Flusight-ensemble | 350.65 | 0.84 | 559.53 | 46 | 90 | 100 |
| SigSci-TSENS | 356.15 | 0.85 | 574.29 | 33 | 90 | 100 |
| CU-ensemble | 363.88 | 0.87 | 552.35 | 15 | 68 | 100 |
| Flusight-baseline | 418.70 | 1.00 | 684.62 | 26 | 88 | 100 |
| UT_FluCast-Voltaire | 440.00 | 1.05 | 633.96 | 38 | 85 | 100 |
| GT-FluFNP | 461.10 | 1.10 | 660.28 | 44 | 75 | 100 |
| CEID-Walk | 471.77 | 1.15 | 679.19 | 28 | 58 | 89 |
| SGroup-RandomForest | 495.62 | 1.18 | 727.54 | 69 | 100 | 100 |
| MOBS-GLEAM_FLUH | 486.49 | 1.22 | 727.21 | 18 | 63 | 94 |
| IEM_Health-FluProject | 613.93 | 1.47 | 785.61 | 42 | 97 | 100 |
| LUcompUncertLab-TEVA | 618.39 | 1.52 | 895.44 | 12 | 56 | 89 |
| LUcompUncertLab-VAR2K | 643.17 | 1.58 | 928.09 | 14 | 61 | 89 |
| UVAFluX-Ensemble | 757.94 | 1.81 | 655.74 | 71 | 94 | 100 |
| LUcompUncertLab-VAR2K_plusCOVID | 747.56 | 1.84 | 888.94 | 6 | 14 | 89 |
| LUcompUncertLab-VAR2_plusCOVID | 754.84 | 1.84 | 865.00 | 7 | 16 | 94 |
| LUcompUncertLab-VAR2 | 863.37 | 2.10 | 1077.69 | 4 | 31 | 94 |
| SGroup-SIkJalpha | 1028.91 | 2.46 | 1543.18 | 15 | 49 | 100 |
| GH-Flusight | 1124.18 | 2.80 | 1141.29 | 0 | 1 | 94 |
| LosAlamos_NAU-CModel_Flu | 1395.41 | 3.33 | 1801.90 | 7 | 15 | 100 |
| **2022-23** | | | | | | |
| MOBS-GLEAM_FLUH | 1509.55 | 0.41 | 2270.89 | 42 | 90 | 100 |
| CMU-TimeSeries | 2043.33 | 0.55 | 3064.96 | 67 | 90 | 100 |
| PSI-DICE | 2087.92 | 0.56 | 2780.90 | 53 | 70 | 100 |
| Flusight-ensemble | 2466.05 | 0.66 | 3439.47 | 56 | 75 | 100 |

| Model | Absolute WIS | Relative WIS | MAE | 50% Coverage (%) | 95% Coverage (%) | % of Forecasts Submitted |
|---|---|---|---|---|---|---|
| CU-ensemble | 2800.38 | 0.70 | 3210.70 | 36 | 51 | 85 |
| MIGHTE-Nsemble | 2666.71 | 0.71 | 3480.24 | 53 | 68 | 96 |
| SigSci-TSENS | 2891.23 | 0.73 | 3972.84 | 57 | 74 | 88 |
| UMass-trends_ensemble | 2914.47 | 0.78 | 3870.83 | 52 | 66 | 100 |
| SGroup-RandomForest | 2869.94 | 0.80 | 3993.83 | 57 | 80 | 96 |
| CEPH-Rtrend_fluH | 2886.69 | 0.81 | 3208.62 | 34 | 58 | 88 |
| GT-FluFNP | 2999.90 | 0.81 | 3516.60 | 58 | 68 | 100 |
| UVAFluX-Ensemble | 3007.81 | 0.81 | 4216.99 | 36 | 67 | 100 |
| UNC_IDD-InfluPaint | 3221.15 | 0.95 | 4092.70 | 36 | 75 | 77 |
| UGA_flucast-OKeeffe | 3466.83 | 0.97 | 4306.36 | 32 | 49 | 96 |
| Flusight-baseline | 3724.31 | 1.00 | 4281.00 | 43 | 59 | 100 |
| NIH-Flu_ARIMA | 3910.19 | 1.03 | 4598.75 | 31 | 100 | 92 |
| ISU_NiemiLab-Flu | 5096.20 | 1.26 | 5870.45 | 18 | 38 | 77 |
| JHU_IDD-CovidSP | 8230.60 | 2.22 | 9494.40 | 32 | 64 | 85 |

The Absolute WIS column refers to the Weighted Interval Score for each model across all fifty states, D.C., and Puerto Rico forecast targets. The Relative WIS compares the WIS value of each model to the Flusight-baseline model. All models with a relative WIS score less than one outperformed the baseline model when evaluated solely upon WIS. 95% and 50% coverage values are provided for the percent of times that reported weekly incidence values were within the 95% or 50% prediction intervals respectively, across all the forecast targets submitted by each team. The percent of forecasts submitted is determined by the number of forecast targets submitted by each team out of all possible forecast targets occurring within the duration of the evaluation period.

When only national forecast targets were considered, two additional models, PSI-DICE and UMass-trends_ensemble, outperformed the FluSight Ensemble in terms of relative WIS in the 2021-22 season. In contrast, one fewer models outperformed the FluSight Ensemble in terms of relative WIS in the 2022-23 season.

For national targets, the CMU-TimeSeries and PSI-DICE model forecasts outperformed the FluSight ensemble forecasts for both seasons in terms of relative WIS scores. Consistent differences in coverage values across models were not observed when comparing all jurisdiction results to national only results.

### Disclaimers

Any use of trade, firm, or product names is for descriptive purposes only and does not imply endorsement by the U.S. Government. The findings and conclusions in this report are those of the authors and do not necessarily represent the views of the Centers for Disease Control and Prevention or the National Institutes of Health.
